# Strengthening access to and confidence in COVID-19 vaccines among equity-deserving populations across Canada: An exploratory qualitative study

**DOI:** 10.1101/2024.03.27.24304984

**Authors:** Kainat Bashir, Mariame O. Ouedraogo, Christoffer Dharma, Mercedes Sobers, Vajini Atukorale, Dane Mauer-Vakil, Anushka Ataullahjan, Shaza A. Fadel, Sara Allin

## Abstract

**Introduction:** There is a need to reflect on the COVID-19 vaccine distribution across Canada and the extent to which they considered equity-deserving populations. This paper examined and compared strategies implemented by six Canadian provinces to increase access and promote the uptake of COVID-19 vaccines among selected priority populations. We also explored the factors that impacted the implementation of these strategies.

**Methods:** In six provinces (Alberta, British Columbia, Manitoba, Nova Scotia, Ontario, and Quebec), we conducted an environmental scan of provincial rollout documents and media sources reporting vaccine distribution among selected priority populations: First Nations, Inuit, and Metis; Black communities; essential workers; people experiencing homelessness; and people with disabilities. We subsequently interviewed 39 key informants to validate the environmental scan results, identify additional strategies to increase COVID-19 vaccine uptake, and uncover perceptions of the facilitators and challenges that influenced the strategies’ implementation.

**Results:** Through the environmental scans and key informant interviews, we identified that provincial health authorities employed a panoply of strategies to overcome geographic, financial, and attitudinal barriers to COVID-19 vaccines experienced by the priority populations. Most provinces implemented walk-in, mobile, and pop-up vaccination clinics, mobilized public and private health workforce, and designed multilingual communication materials. Facilitators in implementing COVID-19 vaccination strategies included fostering inter-governmental cooperation, harmonizing communication efforts, leveraging existing relationships and networks, and ensuring representation and leadership of community partners. Challenges to implementing COVID-19 vaccination strategies included uncoordinated communication efforts, inadequate distribution of vaccines to areas with the greatest need, mistrust in the government and healthcare system, vaccine hesitancy, and lack of cultural competence by vaccine providers.

**Conclusions:** This study highlights the divide between well-intentioned strategies and interventions and the reality of on-the-ground implementation. The findings offer valuable insights and can inform the implementation of strategies to distribute vaccines equitably in future large-scale vaccination efforts in Canada and globally.

## BACKGROUND

The COVID-19 pandemic was a global health emergency of unprecedented nature that resulted in an estimated 6.9 million deaths globally and 52,000 in Canada between January 2020 and May 2023 (1). The rapid spread of the novel coronavirus prompted all high-resource countries to institute a myriad of prevention and control measures and invest massively in developing, procuring, and distributing vaccines (2). With the increased availability of COVID-19 vaccines, calls for vaccine equity within and among countries emerged (3–8). Vaccine equity in the context of the COVID-19 pandemic was seen as an imperative to "end this pandemic, restart economies, and begin to tackle the other great challenges of our time" (4). Based on lessons from the H1N1 pandemic (9, 10), key approaches towards domestic vaccine equity in Canada have been ensuring adequate vaccine procurement from pharmaceutical companies and guaranteeing citizens’ free access to COVID-19 vaccines.

Vaccine equity is highly complex and goes beyond ensuring a sufficient procurement of vaccines and reducing end-user vaccine costs. It is also shaped by factors such as accessibility to vaccines and the confidence in their efficacy and the health actors delivering them (2, 5, 6). Vaccine access and confidence are, in turn, influenced by social and economic conditions like physical environment, culture, and experiences of discrimination, racism, and historical trauma (6). Studies from high-income countries have shown that vaccine hesitancy and poor access to vaccination services are higher among Black, Indigenous, and other minoritized populations than non-racialized people due to historical and current systemic racism and distrust of the health system (6–8). In Canada, specifically, an analysis based on the 2020 Canadian Community Health Survey (CCHS), an ongoing yearly cross-sectional survey conducted by Statistics Canada, found that Canadians who self-identify as off-reserve First Nations, Black, or Arab were less likely to have received at least one dose of a COVID-19 vaccine (11). Another population-based survey in Canada also identified that essential non-healthcare workers had lower intention to receive a COVID-19 vaccine than non-essential workers (12). Evidence from other studies from high-income countries also points to homelessness and disability influencing the acceptance of and access to COVID-19 vaccines (13–15).

Guided by its Ethics, Equity, Feasibility, and Acceptability (EEFA) framework (16), Canada’s National Advisory Committee on Immunization (NACI), an independent advisory committee of experts in immunization and other health-related fields, established preliminary guidance for an equitable distribution of COVID-19 vaccines across the country in the context of limited vaccine supply (17, 18). It defined priority populations for immunization based on their likelihood of severe illness and death and transmitting COVID-19 to high-risk individuals, vital employees, and those in risky environments. In its guidelines, NACI also called for considering pre-existing social and structural inequities, magnifying the vulnerability of some population groups (18). The committee suggested using a mix of interventions and strategies to improve vaccine uptake and confidence in identified priority populations, recognizing that a one-size-fits-all vaccine response would not be effective (17, 18). The provinces and territories began rolling out COVID-19 vaccines to their residents soon after the federal government procured the Pfizer-BioNTech and Moderna vaccines and Health Canada approved their use in December 2020 (19). Provincial and territorial governments could consider NACI guidelines when designing and implementing their vaccination eligibility and distribution plans (20).

Despite establishing equity-centered guidelines and achieving one of the highest vaccination rates in the world, with close to 80.5% of the population having received at least two doses of COVID-19 vaccines as of September 2023 (21), Canada still records vaccination rates that vary significantly by geography, race, ethnicity, and working and living conditions (8). This raises the question of how equity was integrated into the immunization approaches employed across the country and how NACI guidelines were translated into practices in provinces. From examples of vaccination campaigns for the H1N1 pandemic (22) and other major public health concerns like measles (23, 24) and human papillomavirus infection (24, 25), it is well recognized that enhancing vaccine access, overcoming vaccine hesitancy, and ultimately achieving vaccine equity, require strategies and interventions addressing the needs and concerns of various population groups.

In light of the variability in vaccination rates by social and economic conditions, this study aimed to understand and reflect on the COVID-19 vaccine provincial response in Canada and how it reached equity-deserving populations. This study sheds some light on this by assessing the provincial strategies employed to promote and facilitate the uptake of and confidence in the COVID-19 first and second vaccine doses among key populations identified by NACI and the factors influencing their implementation. Our findings can support learning about governments’ roles and processes to achieve equitable vaccine access and confidence, particularly in the context of future emergency and routine mass vaccination efforts in Canada and globally.

## MATERIALS AND METHODS

Based on NACI’s guidelines, populations of interest in this study include First Nations, Métis, and Inuit (FNIM) populations, Black communities, essential workers, people with disabilities, and people experiencing homelessness in six pre-selected provinces (i.e., Alberta, British Columbia, Manitoba, Nova Scotia, Ontario, and Quebec) that make up for approximately 90% of the Canadian population.

Our study consisted of an environmental scan complemented by interviews with key informants who could speak about provincial rollouts of COVID-19 vaccines. We first gathered publicly available information about the strategies taken by selected provinces of interest to deliver the COVID-19 first and second vaccine doses to the populations prioritized by NACI. This was done by means of conducting an environmental scan of provincial documents and news releases, scientific literature, and media sources reporting on vaccine prioritization and distribution among our selected populations. The first environmental scan was completed in June 2021, prior to the start of the interview phase. Informed by the “Reach” dimension of the RE-AIM framework (26), we used a content analysis approach to extract data from the documents around the prioritization of populations and the strategies employed to promote COVID-19 vaccine uptake (Table 1). The data extracted was used to generate provincial summaries of the rollout of COVID-19 vaccines for our populations.

**Table 1.** Questions used to conceptualize the Reach dimension of the RE-AIM framework.

|  |
| --- |
| 1. Was this population prioritized? If so, when? |
| 2. Were there clear communication and justification for the choice of this population? |
| 3. What strategies or interventions, if any, were used to reach or engage with the prioritized population? |
| 4. What strategies or interventions, if any, were used to overcome potential challenges to access the vaccine for those populations, including strategies to improve trust and reduce hesitancy? |

We subsequently interviewed individuals from one of the six provinces who either directly served on COVID-19 vaccine rollout committees, task forces, or advisory panels, or had extensive knowledge of the rollout of COVID-19 vaccines in the provinces to contextualize, validate, and supplement findings from the environmental scan. In advance of their interviews, participants were provided with a summary of the COVID-19 vaccine rollout in their province. During their interview, respondents were asked questions aligned with those displayed in Table 1, capturing the principles that guided the prioritization of populations for COVID-19 vaccination in their provinces and the strategies used to ensure adequate uptake of vaccines by the populations of interest in this study. Participants were also asked to reflect on the provincial strategies and interventions, as well as the factors that influenced the strategies’ implementation. An interpretive descriptive qualitative approach was used to analyze the qualitative interview data collected (27). The timing of and rationale for prioritizing our selected population groups during the rollout of COVID-19 vaccines is the focus of another paper conducted by our research group.

Key informants across six selected Canadian provinces (Alberta, British Columbia, Manitoba, Nova Scotia, Ontario, and Quebec) were invited to participate in the study from November 2021 to April 2022 through email. Potential participants were identified through publicly available information and the research team’s professional networks. An information sheet outlining the study’s objectives and accompanying consent form was shared with the interested informants prior to the interview. No incentives were offered for participation. In some cases, participants’ requests for group interviews (i.e., total of five requests) with their colleagues were accommodated. Semi-structured interviews were led by two research team members (MS and VA) and assisted by CD and KB when needed on Zoom and averaged about 1 hour (interview length ranged from 33 to 85 minutes). Field notes were taken during the interviews. MS and VA are both doctoral students in the Epidemiology program at the University of Toronto who each have training in qualitative methods. They both had an awareness of the health systems in the provinces studied through reading literature and conducting the environmental scan (as described above). At the end of each interview, participants were asked if they had any suggestions about who else to interview to gather additional insights on the strategies used to deliver vaccines to our key populations and their implementation. Interviewees were also invited to provide additional resources on the strategies not captured in our initial environmental scan. Recruitment continued until data saturation was reached and no new themes emerged.

Interviews were audio recorded on Zoom, and transcripts were auto-generated through Zoom’s Audio transcript feature. A research team member listened to the audio and edited for accuracy of the transcript. Key informant names and identifying information were removed. Each key informant was assigned a unique number. Anonymized transcripts were also returned to key informants to be verified. None of the respondents expressed concerns about the information they shared. Participants only edited inaudible sections highlighted by the research team.

The transcripts were imported into NVivo 12 software. Data were analyzed independently by research team members (KB, MO, CD, DM) using inductive thematic analysis. We used an inductive analytic approach as it allowed us to look deeply at the context surrounding the vaccination strategy and implementation. An inductive approach allowed us to analyze our data without preconceived categories or theories and identify patterns, categories, or themes that may not have been considered initially (28, 29).

Creswell’s approach to coding was followed (30, 31). Each transcript was first read, and key ideas or responses that addressed the study objectives were highlighted independently by two research team members. These key ideas or responses were assigned a “code.” Codes were operationally defined to be consistently applied throughout the data. Similar codes were grouped to create broader themes, which became our main units of analysis in this study (31). After coding all the interviews, summaries were developed for each theme by reviewing all data coded. Two team members (KB and MO) completed this, followed by revision and consensus on the themes. The team members involved in this process met and compared their interpretations regularly. All disparities in the coding, categorization, and definitions were discussed, and consensus was sought throughout.

After the key informant interviews, we updated and expanded on the list of strategies gathered through an update of the earlier scan of published literature on the COVID-19 vaccine rollout in Canada to account for additional strategies described by key informants as well as other strategies that we might have missed. The consolidated criteria for reporting qualitative research (COREQ) were used to guide reporting for this study (32) (Supplementary file). We received approval from the University of Toronto ethics committee (REB protocol #28098) to conduct the research in October 2021.

## RESULTS

### Characteristics of the participants

A total of 89 key informants across six Canadian provinces (Alberta, British Columbia, Manitoba, Nova Scotia, Ontario, and Quebec) were invited to participate in this research study. We conducted 25 individual and 6 group interviews (based on interviewees’ preference) with 41 key informants via Zoom (and, in one case, responses to our interview guide were provided via email). Two key informants recused their full interview data because of anonymity concerns, leading to 39 key informants’ data being used in the analysis. Most participants were from Ontario, followed by Nova Scotia and Quebec (Figure 1). A total of 27 key informants (71%) represented an academic, government, or research institution, while 29% represented a community organization. Few informants also served on national vaccine advisory boards. Approximately 90% of participants had experience with communicable disease control and vaccine delivery programs before the COVID-19 pandemic.

**Figure 1.**
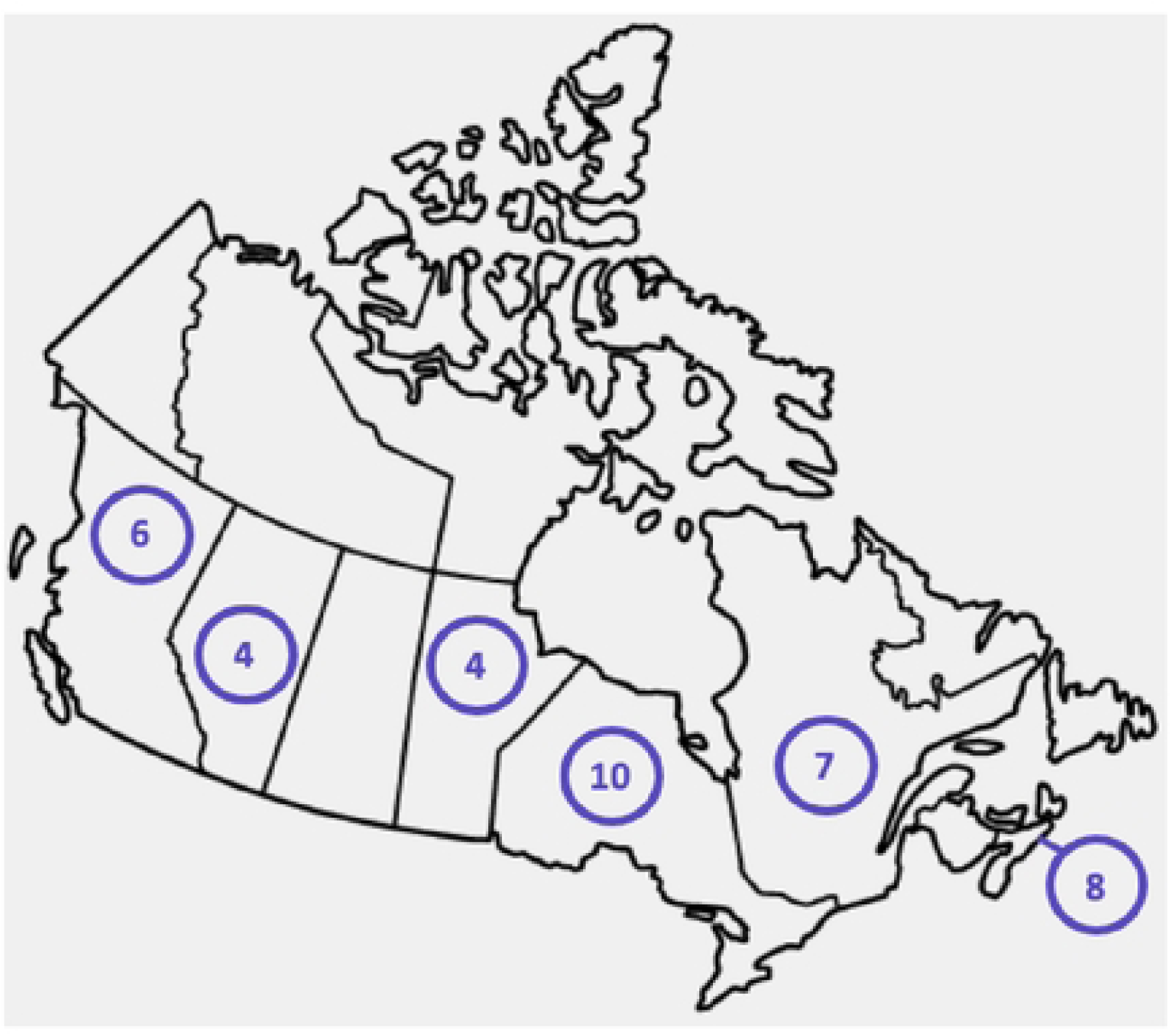
Number of key informants interviewed by province.

### Provincial strategies to improve COVID-19 vaccine uptake by priority populations

Recognizing the need to contain the propagation of COVID-19 cases in the country, provinces utilized various strategies to promote equitable vaccine uptake among FNIM, Black communities, essential workers, people experiencing homelessness, and people with disabilities (Figure 2).

**Figure 2.**
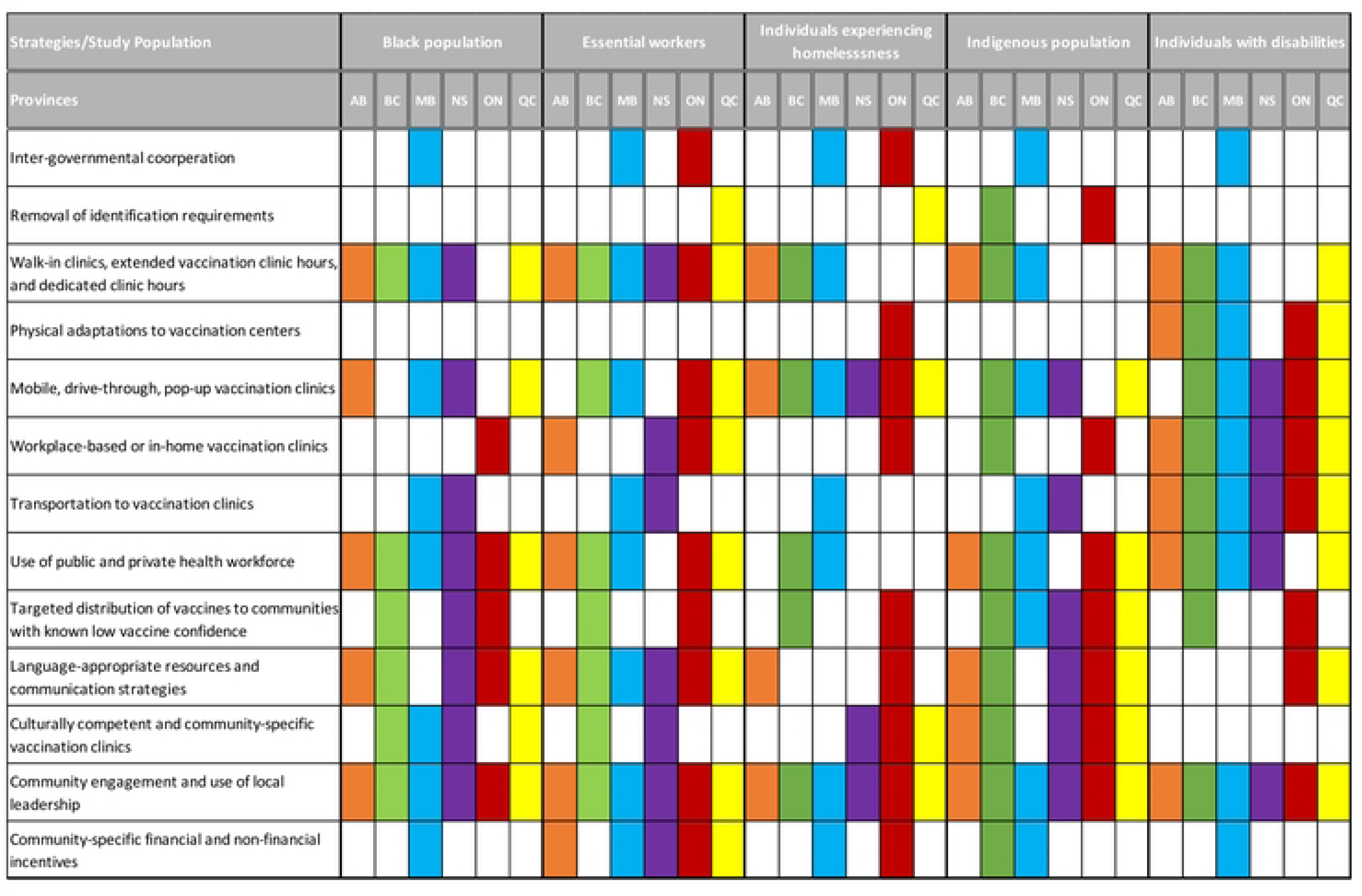
Strategies used to deliver COVID-19 first and second doses to priority populations.

All provinces used the strategy of removing identification requirements to promote equitable access to COVID-19 vaccines. Health insurance cards were not required to book a COVID-19 vaccine appointment to encourage everyone to access COVID-19 vaccines regardless of their residency status or medical insurance coverage. Additionally, in British Columbia and Ontario, key informants noted efforts made at the provincial level to remove the requirement for the Indigenous status card to provide FNIM populations with broader access to COVID-19 vaccines when they were prioritized before the general public.

An array of strategies, including walk-in centres and extended and dedicated vaccination centre hours were implemented in all six provinces to address the accessibility needs of essential workers who could find time to get their vaccinations without difficulty. Specific walk-in vaccination centers were set up for Black communities in some provinces, such as Ontario and Nova Scotia. In other provinces like Manitoba and British Columbia, some clinic hours were dedicated to vaccinating the FNIM populations. Like in some other provinces, in Quebec, efforts were deployed to implement vaccination sites that specifically served people experiencing homelessness. Furthermore, drive-through, mobile, and pop-up vaccination clinics were implemented in several provinces to address the needs of those who found it challenging to find the time or the means to go to the vaccine clinics. As noted by an informant from British Columbia, different strategies were needed to make the vaccines more accessible:

> ***Knowing that there were various groups that might be hesitant, we just made it as possible as we could, using drop-ins, drive-throughs, or outreach or pop-up clinics, we went out to populations, we made it easy for them to come to us. We organized things if they were concerned (…). We did everything possible to make it safe and accessible for them to feel safe about getting a vaccine.*** – Respondent from British Columbia (ID 761)

Most provinces also performed adaptations to their vaccination centres to ease the access and uptake of COVID-19 vaccines by key populations. For example, several respondents indicated that vaccination centres were wheelchair-accessible and low-stimulation areas to serve people with disabilities better.

> ***(We made) sure that our clinics were disabled-appropriate and friendly, that we had separate channels for people who were physically disabled, and that they had places to sit and be comfortable. We worked with advocacy groups to make sure that they felt their needs were being met, so it was very interactive.*** – Respondent from British Columbia (BC01)

In Ontario, there was evidence of efforts made to encourage vaccine access among people experiencing homelessness by providing meals and a safe place for them to socialize.

> ***We were providing meals. We (wanted to) make sure that we were able to provide a warm, welcoming place and didn’t kick them (people experiencing homelessness) out after getting a vaccine, so (we had a) very different clinic style than our normal or generic sort of vaccine clinic*** – Respondent from Ontario (ID 373)

Another strategy to facilitate vaccine access for essential workers involved collaborating with private and government-owned enterprises to set up workplace vaccination centres. The strategy was used in several provinces (Quebec, Alberta, Ontario, and Nova Scotia) to enable workers’ and their families’ access to vaccines in convenient and accessible locations that would not necessitate them to take time off from work. The ability to travel to vaccination sites was especially challenging for people with disabilities, prompting all provinces to initiate transportation services for this sub-population. Furthermore, for people experiencing various disabilities or transportation-related challenges, several provinces organized at-home vaccination efforts. For other key populations, some efforts to offer transportation services were reported in Manitoba and Nova Scotia.

All provinces relied, to varying degrees, on local pharmacies and primary care physicians to administer the first and second doses of COVID-19 vaccines. Involving pharmacies and primary care physicians was seen as an effective way to quickly increase equitable access to vaccines across all population groups, including priority populations. Individuals could get their COVID-19 vaccine doses in an environment they trusted and were comfortable in. Respondents also explained that using pharmacies and primary care physicians was seen to be appropriate for people with mobility or distance-related issues to receive their vaccine at a familiar, convenient, and accessible place:

> ***Having vaccination in local pharmacies increased accessibility but also the confidence (of the) individuals who went to those pharmacies regularly. They have more of a trusting bond with their pharmacists. (…) It’s more personalized and closer to home.*** - Respondent from Quebec (ID 509)

Strategies were also used by provinces to improve the distribution of the first and second doses of the COVID-19 vaccines to communities disproportionately hit by the COVID-19 pandemic and with known low vaccine confidence. One such strategy was the ‘high-priority community strategy’ utilized by Ontario’s government. The strategy consisted of funding community organizations to implement appropriate COVID-19 vaccination initiatives in Ontario’s racialized, low-income, and marginalized communities. The province of British Columbia also adopted a similar strategy, identifying and prioritizing high-transmission communities for vaccination in April 2021. Through the ‘high-transmission neighborhood vaccine plan,’ British Columbians born in 1981 or before (40+) living in 13 communities were prioritized for the AstraZeneca vaccine ahead of the general public.

> ***The High Priority Community Strategy focused on marginalized and racialized populations. We led agencies within those communities that were funded to provide engagement, education, and culturally appropriate support in languages that were particular in those communities. (…) You would see pop-ups at mosques, temples, community centers, and churches. So, very much focused again on particular congregations and community groups. (…) We worked to provide vaccinators who were from those communities, so there was an additional comfort level.*** – Respondent from Ontario (ID 624)

Another approach that informants from Ontario described was the ‘ring-fencing’ approach, which was utilized to allocate a specific number of doses of vaccines for Black populations in the province, as recommended by the province’s Black Scientists’ Taskforce on Vaccine Equity. The concept of “ring-fencing” originates in the fields of business or finance and refers to the act of setting aside a portion of the assets a company owns without necessarily being operated as a separate entity. In Manitoba, informants referred to the ‘push and pull method” that was applied in providing vaccines to FNIM populations living on reserves where the supply of vaccines was controlled based on the community’s needs at the time. During the initial stages of the vaccine rollout, a “push” delivery system was used where FNIM communities received more doses to address the high demand for vaccines. Due to the unpredictable nature of the delivery system at that time, a shift to a more routine “pull” vaccine ordering system was made when the need was met, and high rates were achieved in those communities to ensure that enough doses were available to be administered equitably. Informants in British Columbia described a collaboration between the province of British Columbia and the First Nations Health Authority to implement a “whole community approach” to vaccinate First Nations, regardless of whether they lived in urban areas or on reserve.

> ***(The Whole Community Approach) was quite philosophically different (from how) we looked at the allocation of resources and the rollout. I think what made a huge difference here was looking at and embracing the social determinants of health. And the context by which many (First Nations communities) are living, be it rural, remote, or urban, and the impact on their health and wellness as a result of it (…) We were successful in many regards for advocating for a Whole Community Approach that really honored the lived experience of (the First Nations communities) community.*** – Respondent from British Columbia (ID 370)

All provinces provided resources related to COVID-19 vaccination in multiple languages to ensure that all residents were able to understand the public health messaging, and as a result felt encouraged to book their vaccine appointments. Additionally, provinces supported community-led and culturally-appropriate vaccination centres. Clinics that were specific to the needs of the community and staffed with health professionals from the community were set up, allowing members of the community to feel safe while receiving their COVID-19 vaccine. In some provinces, translators were offered at vaccination sites. Another example of this was capitalizing on the existing relationship several provinces had with Friendship Centres to partner in the delivery of vaccines and address vaccine hesitancy in a safe environment among urban FNIM populations.

Finally, several provinces provided financial and non-financial incentives to increase the uptake of COVID-19 vaccines in priority populations. For example, essential workers in most provinces were offered paid time off to get vaccines. In Ontario, small cash honorariums, coffee gift cards, and meals were used as strategies to encourage vaccination among people experiencing homelessness. Other monetary and non-monetary incentives not specific to communities were used in other provinces (e.g., gift card program in Alberta and lotteries with cash prizes in Manitoba and Quebec).

### Facilitators of the implementation of strategies to improve COVID-19 vaccine uptake by priority populations

Through our analysis of the key informant interviews, we identified the following factors as facilitators of the implementation of provincial strategies used to deliver COVID-19 first and second doses to priority populations: intergovernmental cooperation; harmonized communication efforts; leveraging existing relationships, networks, and infrastructures; and representation and leadership of community partners (Figure 3). While there may be some overlap in these approaches, we will outline distinct examples of each below.

**Figure 3.**
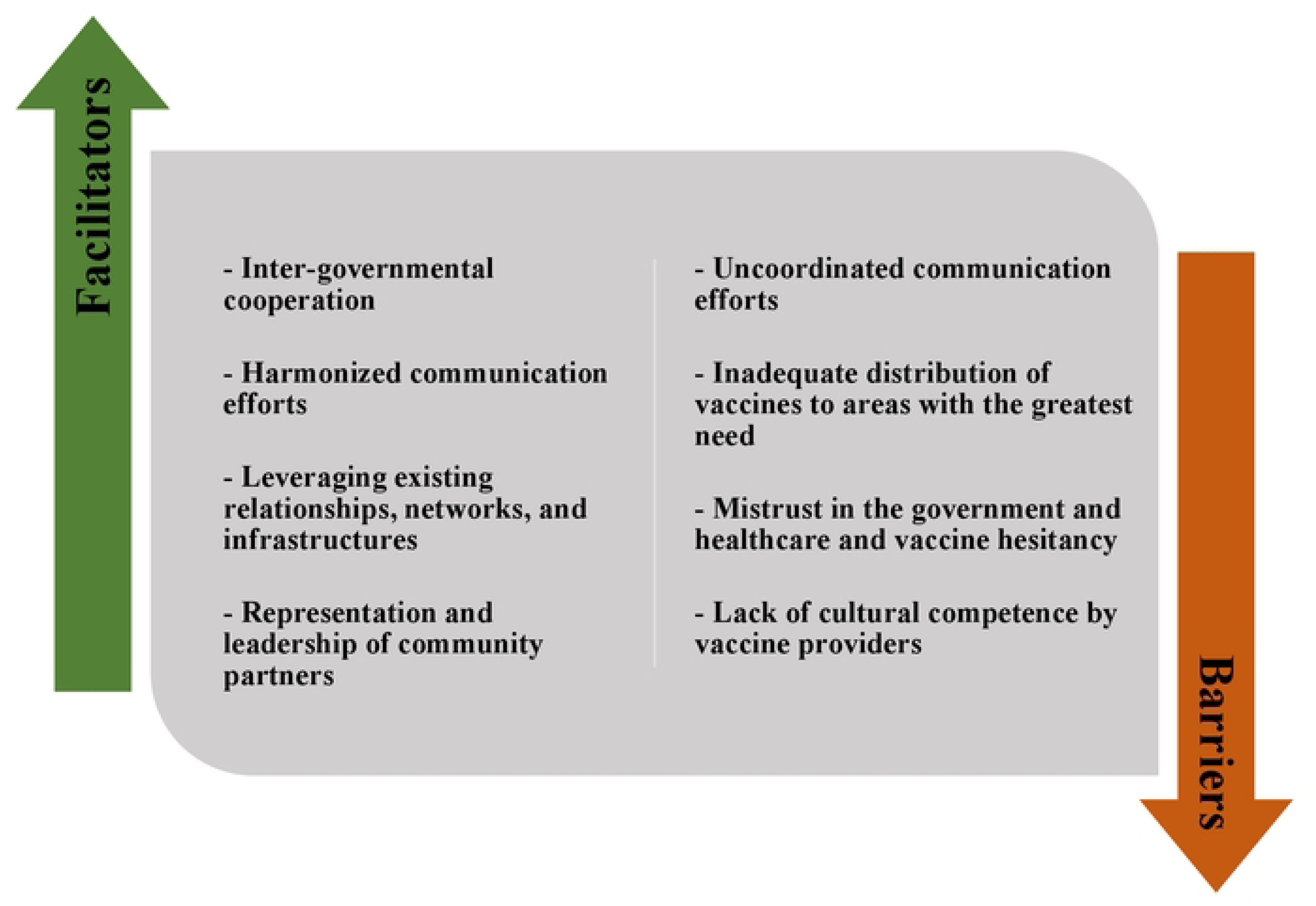
Facilitators and challenges to the implementation of provincial strategies used to deliver COVID-19 first and second doses to priority populations.

#### Inter-governmental cooperation

Several provinces adopted a whole-of-government approach, calling for the cooperation of all government departments and agencies to foster the operationalization of the COVID-19 vaccine uptake strategies. Provinces collaborated with government leadership at the local level and with the federal government, allowing the implementation of their strategies in a more integrated, collaborative, inclusive, and efficient way, thus minimizing gaps in the COVID-19 vaccine rollout.

> ***I think one of the critical success factors in Manitoba was that kind of collective, cooperative model. All hands-on deck, all jurisdictions working together (…) As much as possible de-politicized and focused on a healthcare response.*** – Respondent from Manitoba (ID 632)

#### Harmonized communication efforts

Some participants expressed that their provincial vaccine communication tools and resources were clear, straightforward, uniform across jurisdictions, constantly updated, and delivered by trusted public health experts, contributing to raising the confidence and building the trust of communities in the provinces’ COVID-19 vaccine rollout. Communication materials were shared with all health authorities as well as media outlets to ensure coordination and harmonization in the messaging regarding COVID-19 vaccines, which ultimately promoted the uptake of COVID-19 vaccines in several communities.

#### Leveraging existing relationships, networks, and infrastructures

Leveraging existing relationships, networks, and infrastructures across organizations was a significant determinant that drove the various strategies employed by the provinces to improve vaccine uptake among key populations. An example of this was the pre-existing relationship between the provincial government of Alberta, the Metis Nation of Alberta, and the Metis General Counsel of Alberta. According to a key informant in Alberta (ID 672), they could ***“build on past relationships, past infrastructure, and leveraging them.”*** These pre-existing partnerships provided a baseline of trust and credibility with the Metis communities of Alberta, which helped with rolling out the vaccines. They also allowed for the delivery of vaccines in safe environments for Metis and other Indigenous Peoples in the province. Other examples include provinces capitalizing on their connections with disability-focused advocacy groups and shelter networks serving people experiencing homelessness to identify the best ways to deliver vaccines effectively.

#### Representation and leadership of community partners

Representation and meaningful engagement of community- and faith-based organizations in the decision-making process were cited as important facilitators of implementing COVID-19 vaccine uptake strategies in all provinces. As expressed by many respondents, by ensuring the representation and leadership of community organizations at all stages, provincial health authorities were able to get diverse perspectives on the COVID-related needs of communities, identify the most suitable mechanisms and approaches to reach individuals with COVID-19 vaccines and address quickly issues arising during the rollout. Some respondents also indicated that the use of language- and community-appropriate communication tools was facilitated by the meaningful involvement of the community in designing those tools.

> ***One of the things that, I think, was done well was to bring a First Nations health expert and a BIPOC health expert to represent the decisions that were made and to give them power […] to make decisions for their own people. I don’t see how we would have had anything close to as much success without handing the real power of decision-making to those individuals.*** – Respondent from Manitoba (ID 698)

Respondents acknowledged that colonial history, vaccine hesitancy, and mistrust were barriers to accepting COVID-19 vaccines in some communities. Accordingly, provincial health authorities worked with community organizations to overcome these issues and ensure adequate and equitable distribution and uptake of first and second doses of COVID-19 vaccines.

> ***Because of the community knowledge we had around the table, we could identify friendly, familiar locations across the province where we were advised that African Nova Scotians were already familiar with or would use. So those may be churches. Sometimes, they were community recreation centers or some other community center. This was all through a lens of acknowledging vaccine mistrust. So, our partners in the African Nova Scotian community really were educational for us with respect to vaccine hesitancy. This is rooted in mistrust. So, we acknowledged and tried to respond.*** – Respondent from Nova Scotia (ID 765)

Through funding structures for community-based organizations, some provinces were able to provide flexible funding to community organizations, which also played a major role in facilitating the distribution of vaccines. It allowed for leadership within communities to exercise more agency over the distribution of vaccines. According to one key informant in Ontario (ID 811), it was important that the government was able to identify the appropriate leadership, provide them with funding, and then step away as ***"government will never be able to move at the pace that (community organizations) do."***

### Challenges to the implementation of strategies to improve COVID-19 vaccine uptake by priority populations

Across the six provinces, key informants described several challenges to the implementation of provincial strategies used to deliver COVID-19 first and second doses to priority populations: uncoordinated communication efforts; inadequate distribution of vaccines to areas with the greatest need; mistrust against government and healthcare and vaccine hesitancy; and lack of cultural competency and institutional racism (Figure 3). While there may be some overlap in these factors, we will outline examples of each below.

#### Uncoordinated communication efforts

While positive aspects were reported on the communication front, some respondents expressed that the lack of clear communication often obstructed the rollout of COVID-19 vaccines to specific communities. For example, health insurance cards were often requested in vaccination centres, although they were not required in any province. Another example reported by some informants was regarding the eligibility of FNIM populations in British Columbia for the vaccines not being communicated across the health authorities, resulting in delays in delivering vaccines to FNIM Peoples.

> ***Some (vaccine administrators) didn’t understand that if somebody showed up and they were younger than the age group (being prioritized at the time), it was okay if they were First Nations (or) Metis. (…) They were not supposed to (ask for ID). There was some confusion, and people didn’t know how to address that issue*** – Respondent from British Columbia (ID 344)

Respondents also noted barriers to quickly getting the provincial multilingual and community-specific vaccine communication out. Individuals faced delays in knowing their eligibility for the vaccines and the closest locations where they could get them.

> ***There were challenges (in communication). We heard people say, ‘Oh, I didn’t even know I was eligible’ or ‘Oh, I didn’t even know (the vaccine) was given there,’ even after people had been eligible for vaccination for weeks. So, there were these barriers, and those doing door-to-door and those in touch with communities who (made the difference and) reached those not necessarily (part of) the majority watching the press conferences.*** – Respondent from Quebec (ID 892)

#### Inadequate distribution of vaccines to areas with the greatest need

The initial sub-optimal distribution of vaccines to areas with the greatest need was also identified as a barrier to implementing COVID-19 vaccine rollout strategies. Some participants alluded to the initial selection of pharmacies preventing the delivery of vaccines to the most-impacted communities.

> ***I remember going to the map (to) see where [the pharmacies offering COVID-19 vaccines were]. It was interesting. They picked two pharmacies across the street from each other; why didn’t they pick (pharmacies in) the hardest-hit areas of Toronto? (…) It’s not always easy to get the pharmacies in the areas that you want. You have to ask; some (pharmacies) say yes, and some say no. Pharmacies aren’t all built from the same cloth and don’t all have the same interest in doing all of this."*** – Respondent from Ontario (ID 436)

#### Mistrust in government and healthcare and vaccine hesitancy

As described by several respondents, people experiencing homelessness often expressed mistrust toward government-led initiatives, which impacted the delivery of vaccines to this population. This was also noted to be an important barrier when it came to the FNIM and Black populations, as they have historically been discriminated against and marginalized by the healthcare system. Furthermore, vaccine hesitancy was described as an intricate issue rooted in a lack of trust in the public health system and government authorities.

> ***We made sure (to connect) with friendship centers, which is where a lot of folks experiencing homelessness will go, and made sure that we were sharing any of our communications and any of kind of like our vaccine opportunities with those places…But honestly, I don’t know what the uptake of that was. Because (…) homeless folks (…) they have certain people they’re okay working with. But they’re not necessarily trusting folks that just swoop in. And so, there’s a lot of those trust issues with that community that are hard to unpack.”*** – Respondent from Alberta (ID 770)

#### Lack of cultural competency and institutional racism

Some participants also indicated how systematic racism reinforced specific groups’ hesitancy toward the COVID-19 vaccines and resulted in an inadequate uptake of COVID-19 strategies such as walk-in and pop-up clinics. Moreover, the lack of consideration for cultural safety in vaccination strategies was perceived as an important barrier to the success of COVID-19 vaccination strategies.

> ***For the large majority of First Nations, Inuit, and Métis People who live in cities, having hospitals implement the vaccines to start with was the worst possible choice for Indigenous Peoples because they don’t feel safe in hospitals. We anticipate that they may experience life-threatening racism in hospitals. And we saw that. We saw people getting pulled out of lines.*** – Respondent from Ontario (ID 767)

## DISCUSSION

This study examined the strategies used by the provincial authorities of six Canadian jurisdictions to expand COVID-19 vaccination uptake and confidence in populations recommended for vaccine prioritization by NACI (i.e., FNIM, Black communities, essential workers, people experiencing homelessness, and people with disabilities). While research into COVID-19 vaccine equity in Canada has gained great momentum (3, 33, 34), the scientific literature documenting the strategies and interventions for equity in COVID-19 vaccination and the factors influencing their implementation remains scarce, as indicated in a recent scoping review (35). Our work, therefore, makes an important contribution to this knowledge gap. Examining this topic through the lens of implementation methodologies and frameworks enabled us to render rich findings and emerging lessons that can guide future governmental efforts to design and implement vaccine rollout strategies that effectively improve vaccine uptake and confidence of diverse equity-deserving populations.

As the supply and availability of COVID-19 vaccines improved in March 2021, provinces demonstrated ingenuity, agility, and innovation in the strategies employed to improve COVID-19 vaccine uptake equitably. Similar to a past study that focused on provincial and territorial preventive and control measures against the spread of COVID-19 (19), we have found nuances and variability in the vaccine strategies employed by provinces. This was expected, as Canada is a decentralized federation, where provinces hold the responsibility of designing and delivering health services, including immunization services, to their populations (19, 20). Studies have reported that a decentralized vaccination approach has put provinces in a position where they could "innovate and mobilize quickly" (36) and reach high vaccination rates (33). Our findings also point to the fact that decentralization encouraged the incorporation of equity into vaccination efforts. However, we also found that vaccine equity was not consistently prioritized in the provinces. Existing literature also noted unintended consequences of decentralization, such as Canadian provinces considering equity to varying extents in their vaccination rollout plans and having limited opportunities for cross-province learning (37, 38).

The findings outline a panoply of strategies used by the six jurisdictions, ranging from removing health card requirements to delivering home vaccination appointments to people with disabilities and offering monetary and non-monetary incentives to people experiencing homelessness. We also found that combinations of strategies were used to address simultaneously vaccine access and vaccine hesitancy. For instance, some provinces like Manitoba and Nova Scotia coordinated transportation to culturally safe and community-specific vaccination clinics for Black and FNIM Peoples. Other provinces like Quebec offered workplace vaccination for essential workers and designed multilingual communication materials. For the success of vaccination efforts, combining interventions enhancing access to immunization services while building the public’s trust has also been discussed in other studies targeting Black populations (39) and other marginalized groups (40, 41).

Through key informant interviews with individuals involved in leading or advising the vaccine rollout, we uncovered several important facilitators and challenges to implementing COVID-19 vaccination strategies in Canada. Numerous success stories shared by the key informants centered on communicating the vaccine strategies used by each provincial authority. Many interviewees saw the provincial vaccine communication approach as transparent and clear, which positively influenced the public’s trust in the vaccination strategies. However, other respondents indicated how uncoordinated communication affected vaccine access to minority groups. A study in Ontario also reported delays in communicating essential information to frontline vaccine providers, which fueled confusion and misunderstanding and disrupted vaccination efforts for diverse communities (42).

Our findings also align with previous research emphasizing the importance of working with priority groups to deliver COVID-19 vaccine strategies effectively (43). Partnerships between the government and community-based organizations enabled provincial authorities to recognize and address pre-existing social, economic, and structural barriers various communities face during vaccination implementation. By allocating flexible funding to community-based organizations, provinces also provided opportunities to implement culturally adapted vaccination services. Still, as raised by many interviewees, community involvement could have been strengthened throughout the rollout of COVID-19 vaccines to overcome barriers, such as the lack of cultural safety, vaccine hesitancy, and mistrust in public health authorities. Other published studies have reported similar conclusions suggesting that provinces engage communities tardily and unevenly, exacerbating vaccine barriers for equity-deserving communities (13, 33, 37).

We also found that leveraging existing infrastructures was an important positive influence in implementing vaccine strategies. For instance, the use of existing shelters and points of gathering maximized vaccine access among people experiencing homelessness. Similar findings were observed by enabling essential workers to get vaccinated in their place of work. These results also align with published articles (14, 44). Still, our study also identified that the inadequate distribution of infrastructure across neighborhoods requires explicit attention to guarantee fair vaccine distribution more generally.

### Strengths and limitations

Through this study, we were able to gather a wide variety of perspectives on the provincial strategies to promote equitable vaccine access and uptake of COVID-19 vaccines to equity-deserving populations of Canada. Our work sheds light on a topic that has received little attention despite the importance of appraising sub-national variability in approaches and determinants for equitable mass emergency and routine vaccination efforts in the future.

Still, we acknowledge certain limitations. First, we limited our study to six provinces, meaning we missed important insights from the excluded provinces and territories. However, our team tried to capture diverse provinces in terms of geography, at-risk populations, and language. In the early planning stages of the study, we also identified variability in the vaccine prioritization of selected key populations among the six provinces (45). A second limitation to note is that we only interviewed a few key informants per province (i.e., between 4 – 10 informants per province); therefore, the perspectives gathered do not represent an entire province.

It is important to mention that although our use of qualitative interviews with key informants and environmental scans aimed to thoroughly identify all the strategies and interventions deployed by the six provinces, we may have missed those not publicly available online or mentioned by our key informants. Still, we tried to mitigate this limitation by regularly updating our environmental scans throughout this study. This study also does not capture the community-based initiatives known to have filled the gaps in provincial efforts (46). We were also unable to explore the degree to which similar strategies were implemented across the six provinces and how they impacted vaccine acceptance and uptake in priority populations.

Additionally, it would have been important to co-develop this research with community-level members of the priority groups or organizations partnering with government and public officials to reach priority populations. While key informants interviewed shared their reflections and opinions on the effectiveness and appropriateness of the strategies identified, it is important to note that their personal experiences and viewpoints do not necessarily reflect those of members of the priority populations. Assessing whether the strategies adequately addressed our priority populations’ needs was beyond this project’s scope. Still, some research has already emerged on the fact that strategies, despite best intentions, are needed to respond more adequately to the needs of specific communities (13).

Future research should expand this study’s findings by measuring other domains of the RE-AIM framework (i.e., effectiveness, adoption, and implementation) as well as other key implementation outcomes, such as the acceptability of the various strategies in responding to the needs of diverse priority groups. This study was conducted in the early stages of the vaccine rollout when the context was too dynamic, and the availability of secondary data was too limited to assess the other dimensions of the RE-AIM framework. The involvement of priority populations in co-developing and co-interpreting future research endeavors will be important.

## CONCLUSIONS

Promoting equitable distribution of vaccines is crucial in minimizing adverse health effects and attaining widespread immunity. Our findings contribute to the growing literature on approaches for equitable, efficient, and effective vaccine distribution in Canada and beyond. Understanding the facilitators of and challenges to implementing vaccine rollout strategies will also better guide government actors in their implementation, funding, and partnership efforts to improve vaccine uptake and confidence for high-risk and equity-deserving communities moving forward. We recommend that government actors and policymakers pay closer attention not only to the structural barriers to vaccines faced by diverse communities but also to the importance of greater representation and engagement of communities and clear and consistent communication during the deployment of vaccine rollout strategies. Accounting for these recommendations will help promote equitable vaccine distribution in forthcoming large-scale emergency or routine vaccination programs within Canada and globally.

## Data Availability

The manuscript and its supporting information files include all relevant data from the environmental scans. Data from the qualitative interviews cannot be shared publicly because they contain potentially identifiable information on the study’s participants. Data may be available upon reasonable request by the University of Toronto Research Ethics Board.

## ACKNOWLEDGMENTS

The study team acknowledges the interviewees for their time and participation during this study. We also acknowledge the students and professors in the Implementation Science Trainee Cluster at the Dalla Lana School of Public Health, University of Toronto, whose perspectives helped shape this work.

## Notes

### Competing Interest Statement

The authors have declared no competing interest.

### Author Declarations

We received approval from the University of Toronto ethics committee (REB protocol #28098) to conduct the research.

